# Causal relationship between inflammatory cytokines, metabolites and arrhythmia: a mendelian randomization study

**DOI:** 10.1101/2024.11.29.24318214

**Authors:** Yu-fei Xie, Ling-hui Tang, Feng Huang, Zhi-yu Zeng

## Abstract

**Background:** This study aims to explore the causal relationships between inflammatory cytokines (ICs), metabolites, and the risk of arrhythmia through Mendelian Randomization (MR) analysis.

**Methods:** The causal associations were analyzed using five different MR analysis methods. Additionally, reverse MR analysis was performed to assess the impact of arrhythmias on these ICs and their metabolites.

**Results:** The MR analysis revealed that Oncostatin-M receptor (OSM) was significantly associated with an increased risk of arrhythmia (OR = 1.0812, p < 0.05), along with other ICs such as CXCL11 (OR = 1.0586), SIRT2 (OR = 1.0521), and FGF5 (OR = 1.0520). Five were positively correlated with arrhythmia risk, including X-22776 (OR = 1.071, p = 0.022) and tricosanoylsphingomyelin (OR = 1.066, p = 0.035).Mediation analysis demonstrated that FGF5 influences arrhythmia risk through its metabolite 1-palmitoyl-2-oleoyl-GPE, with a mediated effect accounting for 5.1% of the total effect.

**Conclusions:** Our findings suggest that specific ICs and metabolites contribute to the pathogenesis of arrhythmia. In particular, FGF5 and its metabolite 1-palmitoyl-2-oleoyl-GPE are implicated in increased arrhythmia risk, highlighting potential metabolic targets for therapeutic intervention.

## Introduction

Arrhythmias represent a group of conditions characterized by irregular heart rhythms, which can range from harmless to life-threatening. Globally, arrhythmias affect millions of people and are a leading cause of morbidity and mortality[1]. Atrial fibrillation, one of the most prevalent forms of arrhythmia, is associated with an increased risk of stroke, heart failure, and sudden cardiac death [2]. The pathogenesis of arrhythmias is multifactorial, involving a complex interplay of genetic, structural, and environmental factors that lead to abnormal electrical conduction in the heart [3]. Current treatment modalities, including antiarrhythmic drugs, catheter ablation, and implantable devices, provide symptomatic relief and reduce the risk of adverse events but have limitations such as high recurrence rates and potential side effects [4].

Recent research has highlighted the role of inflammation and metabolic dysregulation in the development and progression of arrhythmias. ICs such as IL-6 and tumor necrosis factor-alpha (TNF-α) have been implicated in cardiac electrical and structural remodeling, which can predispose individuals to arrhythmic events[5, 6]. These cytokines can alter ion channel function, disrupt calcium handling, and modify the autonomic nervous system, creating an arrhythmogenic substrate[7]. Additionally, metabolites resulting from inflammation-related metabolic pathways have been shown to influence arrhythmogenesis, suggesting a potential link between metabolic disturbances and arrhythmic risk [8]. Despite these associations, the precise causal relationships between specific ICs, metabolites, and arrhythmia risk remain poorly understood. Traditional observational studies are often limited by confounding factors and reverse causality, necessitating the use of more robust methods to elucidate these complex interactions.

Mendelian Randomization (MR) offers a powerful tool to explore the causal pathways between exposures (e.g., ICs and metabolites) and outcomes (e.g., arrhythmia) by using genetic variants as instrumental variables[9]. MR can overcome some of the limitations of observational studies, reducing confounding and providing more reliable evidence of causality[10]. Previous MR studies have elucidated causal roles of cytokines in cardiovascular diseases, but their relationship with arrhythmias requires further exploration[11]. This study employs a two-sample MR approach to investigate the causal effects of key ICs and their metabolites on arrhythmia risk. By examining these mediating mechanisms, this study aims to provide novel insights into arrhythmogenesis and identify potential therapeutic targets.

## Materials and methods

### Data sources

The summary statistics for ICs were collected from (https://www.phpc.cam.ac.uk/ceu/proteins), which included 91 ICs(Ebi-a-GCST90274758 to Ebi-a-GCST90274848).[12] Data for 1400 metabolites (GCST90199621-GCST90201020) were sourced from the GWAS catalog (Genome-wide association studies),.[13] arrhythmia data were obtained from Finngen_R10_ CARDIAC_ARRHYTM.gz, which includes 74,000 arrhythmia cases and 239,778 controls (https://storage.googleapis.com/finngen-public-data-r10/summary_stats/finngen_R10_CARDIAC_ARRHYTM.gz). All patients in this study were of European descent.

### Study design

In this study, we performed a two-step MR analysis to determine the relationship between ICs and genetically predicted risk of arrhythmia, as well as to explore whether metabolites could mediate this association. Single nucleotide polymorphisms (SNPs) were chosen as instrumental variables (IVs) for MR based on three criteria: 1) a strong association with exposure factors, 2) independence from confounding factors, and 3) a clear relationship with the outcome event. The first step involved using two-sample MR to evaluate the causal effects of ICs and metabolite traits on arrhythmia, identifying those highly associated with arrhythmia risk. The second step assessed the causal impact of the filtered ICs on the filtered metabolite traits and calculated the mediation proportion of each metabolite in the effect of ICs on arrhythmia. Additionally, to ensure no overlap in study subjects, the SNPs representing exposures and outcomes were derived from different research sources. The study design is illustrated in Figure 1.

**Figure 1.**
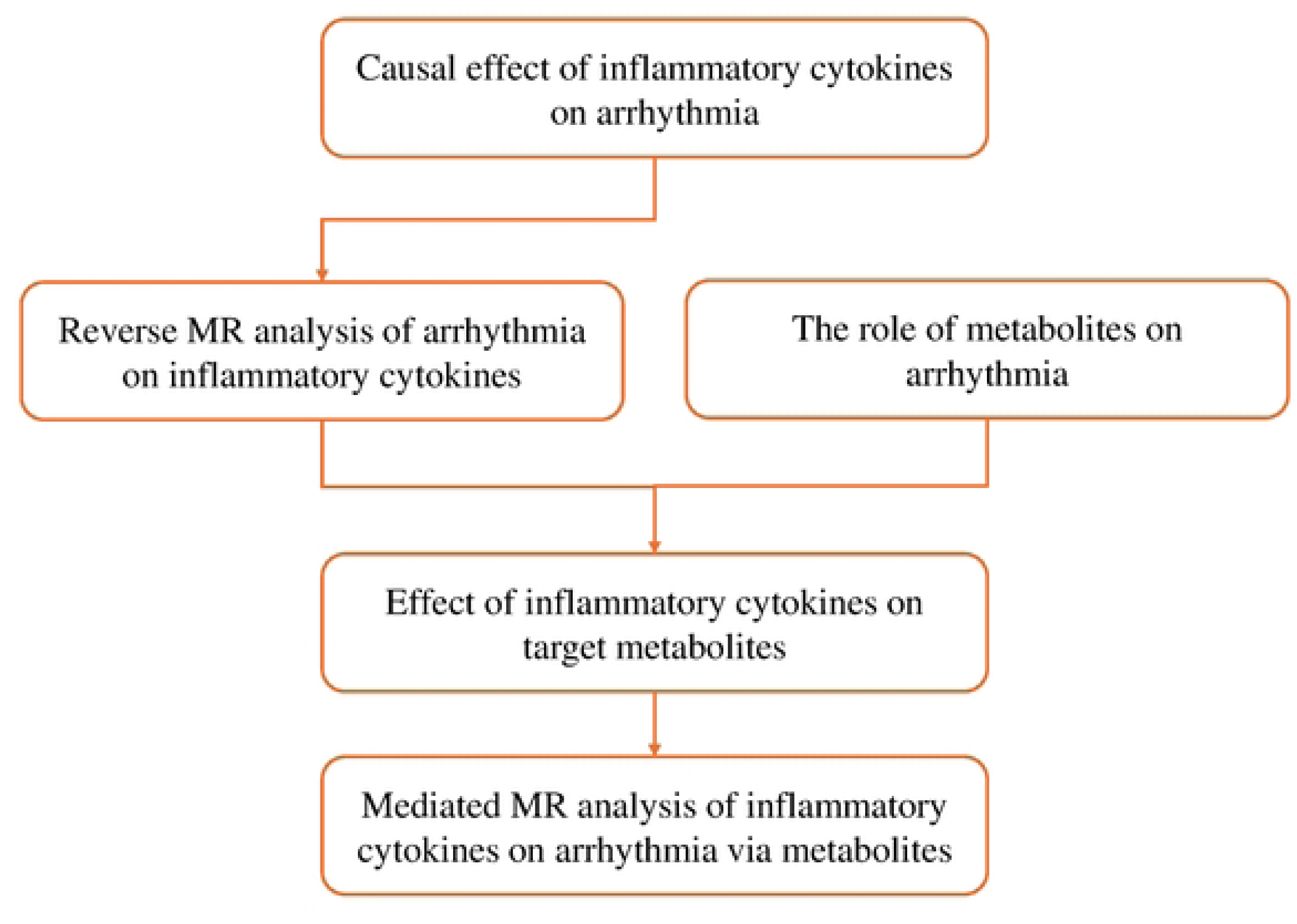

### Genetic IVs selection

The filter condition for SNPs used as IVs for ICs and metabolite traits was set to p < 1×10-5. SNPs used as IVs for arrhythmia were determined using a stricter threshold of p < 5×10-8. We clumped all these genetic variants with a threshold of R² < 0.001 within a 10,000 kb clumping distance. Finally, we applied the F-statistic to screen for SNPs, calculated as β divided by the square of the standard error, with a cut-off value of 10. We then searched the obtained SNPs for potential confounders and bypassing factors (e.g., age, sex, race, other diseases) using the PhenoScannerV2 (http://www.phenoscanner.medschl.cam.ac.uk/).

### Statistical analysis

All statistical analyses were conducted using R version 4.3.3 (https://www.r-project.org/). Two-sample MR analyses were performed utilizing the "Two Sample MR" package, the "Variant Annotation" package, and the "ieugwasr" package. Among the five MR analysis methods ("MR Egger," "Weighted Median," "Inverse Variance Weighted (IVW)," "Simple Mode," and "Weighted Mode"), IVW was used as the primary method for causal estimation due to its higher precision.[10, 14–16] P< 0.05 was deemed to signify a significant association between the exposure and outcome. We utilized the online tool (http://cnsgenomics.com/shiny/mRnd/) to analyze statistical power. Cochran’s Q statistic, based on the IVW and MR Egger methods, was used to assess the level of heterogeneity. To detect pleiotropy, we employed the MR-Egger intercept test and the MR-PRESSO method (Mendelian Randomization Pleiotropy and Outlier), using the "MR-PRESSO" package. MR-PRESSO was also utilized to adjust for horizontal pleiotropy by removing outliers and to assess significant differences before and after this correction. The leave-one-out approach was used to explore the potential impact of individual genetic variants. The “product of coefficients” method was applied to evaluate the indirect effects of ICs on arrhythmia risk via potential mediators, and the δ method was used to calculate the standard error of these indirect effects.

## Results

### Causal effects of ICs on arrhythmia

In this study, 2725, 31908, and 187 SNPs as instrumental variables (IVs) for 91 ICs, 1400 metabolites, and arrhythmia(Tables S1, S2, and S3). Two-sample MR analysis demonstrated a causal relationship between 8 ICs and arrhythmia (Figure 2). Notably, The IVW method indicated that OSM receptor Oncostatin-M levels showed the strongest association with an OR of 1.0812, suggesting a significant relationship with increased risk of arrhythmia. Other factors such as CXCL11 (OR = 1.0586), SIRT2 (OR = 1.0521), and FGF5 (OR = 1.0520)were positively correlated with arrhythmia, potentially promoting inflammation and cardiac destruction. Interestingly, CD40L,GDNF, LIFR and IL-6 showed an OR less than 1 (0.9631,0.9498,0.9387 and 0.9318, respectively), suggesting a possible protective effect against arrhythmia.

**Figure 2.**
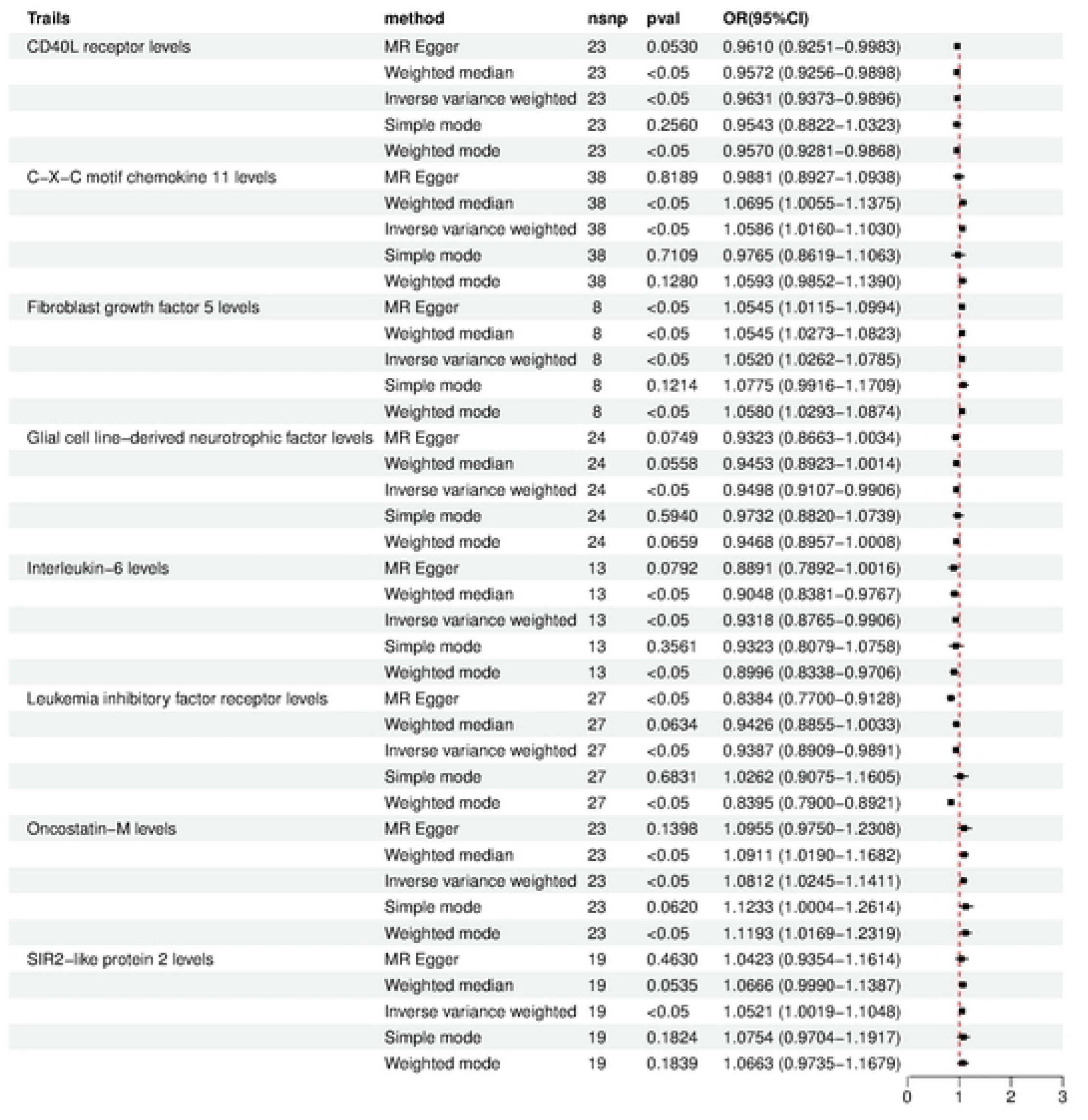

### Reverse MR analysis of ICs by arrhythmia

In the context of MR analysis, we investigated the causal relationship between various ICs and arrhythmias. This approach aimed to identify potential ICs that might either directly contribute to or be influenced by arrhythmic events. For this purpose, we performed a reverse MR analysis to evaluate the causal effects of different ICs on the occurrence of arrhythmias.

The reverse MR analysis indicated no significant causal relationship between the levels of the evaluated ICs and arrhythmias. Specifically, the IVW method indicated that the OSM (OR = 1.001, 95% CI [0.954, 1.071], p = 0.972)and CXCL11 (OR = 1.003, 95% CI [0.960, 1.048], p = 0.899), the results showed no significant association with arrhythmia risk. Similarly, FGF5 known for its involvement in various cellular processes, displayed an odds ratio of 1.078 (95% CI [0.937, 1.106], p = 1.240), indicating no notable effect on arrhythmias. Leukemia inhibitory factor receptor (LIFR) and CD40L receptor levels (CD40L) also showed no significant relationships, with ORs of 1.019( 95% CI [0.939, 1.106], p = 0.648) and 1.027 (95% CI [0.985, 1.071], p = 0.208), respectively. Finally, SIRT2, which has been studied for its role in cellular stress response, did not exhibit a causal association with arrhythmias 1.014 (95% CI [0.972, 1.058], p = 0.521). These findings suggest that the ICs analyzed, including OSM, CXCL11, FGF5, LIFR, CD40L and SIRT2, do not have a direct causal effect on arrhythmias.

### Effects of metabolites on arrhythmia

The IVW method indicated that 98 metabolites, five of these metabolites were found to be positively associated with an increased risk of arrhythmia, suggesting their potential involvement in the pathogenesis of arrhythmia. The strongest association was observed for X-22776 levels, with an OR of 1.071 (95% CI: [1.010, 1.136], p = 0.022). Following this, tricosanoylsphingomyelin (d18:1/23:0) demonstrated a significant association with an OR of 1.066 (95% CI: [1.004, 1.131], p = 0.035). Additionally, palmitoyl sphingomyelin (d18:1/16:0) levels (OR = 1.064, 95% CI: [1.009, 1.123], p = 0.023), cytosine levels (OR = 1.063, 95% CI: [1.008, 1.120], p = 0.023) and lignoceroylsphingomyelin (d18:1/24:0) levels(OR: 1.062,95% CI: [1.001, 1.127],p = 0.046) were also associated with an increased risk of arrhythmia.

In contrast, five metabolites were negatively correlated with arrhythmia, indicating they may offer a protective effect. The level of X-24546 showed the strongest negative correlation (OR = 0.942, 95% CI: [0.917, 0.968], p = 0.014). Other metabolites with an OR of less than 1 included the Phosphate to N-palmitoyl-sphingosine (d18:1 to 16:0) ratio (OR = 0.934, 95% CI: [0.889, 0.981], p = 0.006), X-25828 levels (OR = 0.930, 95% CI: [0.877, 0.987], p = 0.017), P-cresol sulfate levels (OR = 0.927, 95% CI: [0.871, 0.987], p = 0.018), and the 4-methyl-2-oxopentanoate to 3-methyl-2-oxobutyrate ratio (OR = 0.911, 95% CI: [0.858, 0.968], p = 0.002). This dichotomy in OR values high lights the complex role lipid metabolism may play in arrhythmia, with some metabolites potentially increasing disease risk while others may provide protection.

### Effects of ICs on target metabolites

The result shows the results of MR Analysis to arrhythmia the effect of inflammatory cytokine Fibroblast growth factor 5 -arrhythmia on metabolite X-22776 levels. Although other methods showed no significant correlation, IVW method showed significant correlation with an OR of 1.071 (95% CI: [1.010, 1.136], p = 0.022), indicating that cytokines have a potential protective effect on arrhythmia.

### Mediation MR analysis

We analyzed the causal effect of FGF5 on 1-palmitoyl-2-oleoyl-GPE (16:0/18:1) and its effect on cardiac arrhythmias, as demonstrated in Figure 3. Overall, the analysis indicates that FGF5 positively influences the levels of its metabolite, 1-palmitoyl-2-oleoyl-GPE (16:0/18:1), which in turn is associated with an increased risk of developing cardiac arrhythmias. FGF5 shows an OR of 1.065 for its effect on 1-palmitoyl-2-oleoyl-GPE (16:0/18:1) (95% CI: [1.003, 1.132], p = 0.041), while 1-palmitoyl-2-oleoyl-GPE (16:0/18:1) itself has an OR of 1.042 for cardiac arrhythmias (95% CI: [1.014, 1.070], p = 0.003). This suggests a positive regulatory relationship between this inflammatory factor and its metabolite, ultimately contributing to an elevated arrhythmia risk. Finally, we describe in the table the mediating role of 1-palmitoyl-2-oleoyl-GPE (16:0/18:1) in the FGF5 to cardiac arrhythmias causal pathway.

**Figure 3.**
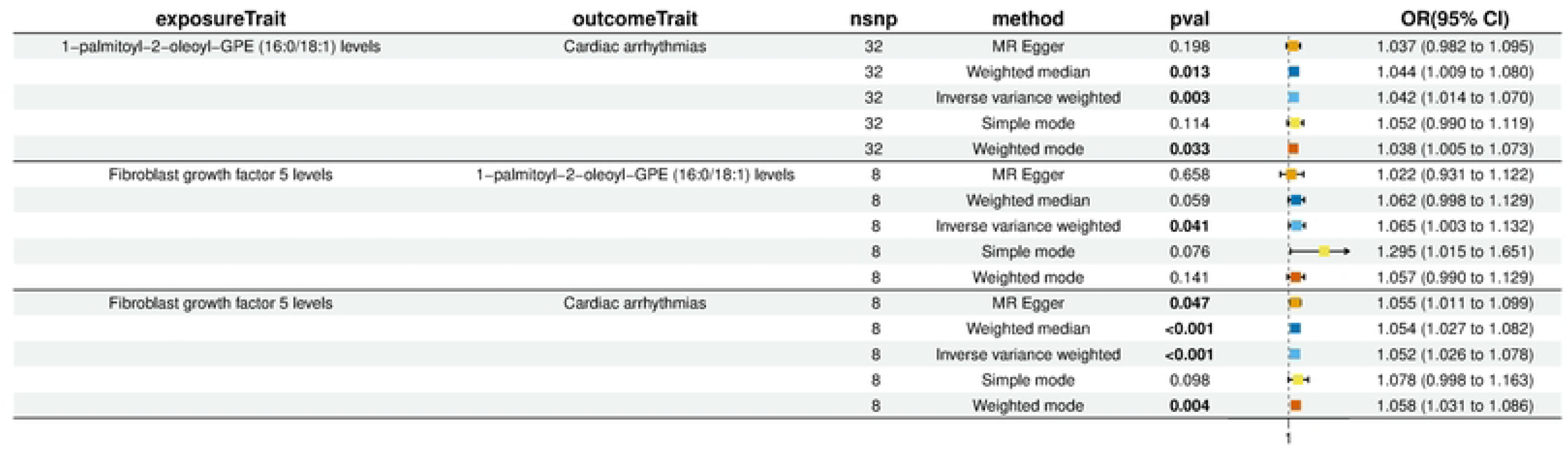

The mediating effect of 1-palmitoyl-2-oleoyl-GPE (16:0/18:1) in the FGF5-cardiac arrhythmias causal pathway was 0.00258 (95% CI: [-0.0014, 0.0066]), accounting for 5.1% of the total effect.

In addition, we explored the direct causal effect of FGF5 on cardiac arrhythmias. The analysis revealed that FGF5 has a significant positive association with cardiac arrhythmias, with an OR of 0.0409(95% CI: [-0.0924, 0.680], p < 0.001). This highlights the complex interplay between FGF5 and its metabolite in the development of cardiac arrhythmias, with both factors contributing to an increased risk. The involvement of 1-palmitoyl-2-oleoyl-GPE (16:0/18:1) as a mediator provides additional insight into how metabolic regulation might influence the pathogenesis of cardiac arrhythmias, supporting the need for further investigation into these molecular mechanisms.

## Discussion

In this MR study, we explored the causal relationships between ICs, metabolites, and arrhythmia. Our findings revealed several significant associations, indicating that specific ICs and metabolites may contribute to the pathogenesis of arrhythmia. Notably, OSM displayed the strongest positive correlation with arrhythmia risk (OR = 1.0812), followed by CXCL11, SIRT2, and FGF5, all of which were also positively associated with inc reased arrhythmia risk. Conversely, CD40L, GDNF, LIFR, and IL-6 exhibited protective effects, suggesting that not all inflammatory pathways uniformly contribute to arrhythmia development. Moreover, five metabolites, including X-22776 and tricosanoylsphingomyelin, were found to increase the risk of arrhythmia, while others, such as X-24546 and the phosphate to N-palmitoyl-sphingosine ratio, appeared to confer protection. Mediation analysis indicated that FGF5 may influence arrhythmia risk through its metabolite, 1-palmitoyl-2-oleoyl-GPE, accounting for 5.1% of the total effect. These findings provide novel insights into the complex interplay between inflammation, metabolism, and arrhythmia.

Our results align with the growing body of evidence that supports the role of inflammation in arrhythmogenesis. Previous research has highlighted the importance of cytokines such as IL-6 and TNF-α in promoting cardiac inflammation, fibrosis, and electrophysiological changes, all of which contribute to arrhythmias [17, 18]. In particular, the association between OSM and arrhythmia risk is consistent with its known role in cardiac remodeling and fibrosis. OSM is a pleiotropic cytokine that mediates various inflammatory responses and has been implicated in the progression of heart failure and fibrosis, both of which are key factors in the development of arrhythmias [19]. The strong association we observed between OSM and arrhythmia suggests that it may serve as a potential therapeutic target for preventing or mitigating arrhythmogenic processes. Similarly, the positive association of CXCL11 with arrhythmia risk is supported by its role in immune cell recruitment and inflammation, processes that are critical in the pathophysiology of cardiac diseases [20, 21]. Our findings regarding the protective effects of certain cytokines, including IL-6 and CD40L, challenge the traditional view that inflammation uniformly promotes arrhythmia. IL-6, a well-known pro-inflammatory cytokine, has been implicated in various cardiovascular diseases, including heart failure and atrial fibrillation (AF) [17]. However, recent studies have suggested that IL-6 may also have context-dependent anti-inflammatory properties, particularly in the regulation of immune responses and tissue repair [22]. Our study’s findings align with this emerging perspective, indicating that IL-6 may exert a protective effect in certain arrhythmogenic contexts. Similarly, CD40L, a cytokine involved in immune cell activation, has been shown to have both pro-and anti-inflammatory roles, depending on the disease setting [23, 24]. Our results suggest that CD40L may protect against arrhythmia by modulating immune responses in a way that reduces inflammation-induced cardiac damage.

The identification of metabolites associated with arrhythmia risk provides further insight into the complex metabolic processes underlying this condition. Our analysis showed that several sphingomyelins, including tricosanoylsphingomyelin and palmitoyl sphingomyelin, were positively correlated with arrhythmia risk. Sphingomyelins are bioactive lipids that play important roles in cell membrane structure, signaling, and inflammation [25]. Elevated levels of sphingomyelins have been linked to increased cardiovascular risk, particularly in the context of atherosclerosis and myocardial infarction [18]. The positive association between sphingomyelins and arrhythmia in our study is consistent with these findings, suggesting that altered lipid metabolism may contribute to arrhythmogenic processes. On the other hand, metabolites such as X-24546 and the phosphate to N-palmitoyl-sphingosine ratio were negatively associated with arrhythmia, indicating their potential protective roles. These findings highlight the dual nature of lipid metabolism in cardiovascular diseases, where certain lipid species may promote disease progression, while others may offer protection by modulating inflammation and oxidative stress [26].

Our mediation analysis further elucidated the role of FGF5 and its metabolite, 1-palmitoyl-2-oleoyl-GPE, in arrhythmogenesis. FGF5 is a member of the fibroblast growth factor family, which is involved in a wide range of cellular processes, including cell growth, differentiation, and angiogenesis [27]. Previous studies have implicated FGF5 in the regulation of vascular remodeling and smooth muscle cell proliferation, both of which are critical processes in the development of cardiovascular diseases [27, 28]. Our study adds to this body of knowledge by demonstrating that FGF5 influences arrhythmia risk through its effect on 1-palmitoyl-2-oleoyl-GPE, a metabolite involved in lipid metabolism. This finding underscores the importance of metabolic regulation in the inflammatory cascade leading to arrhythmia and suggests that targeting specific metabolic pathways may offer novel therapeutic approaches for reducing arrhythmia risk. The mediating role of 1-palmitoyl-2-oleoyl-GPE, accounting for 5.1% of the total effect, further supports the hypothesis that metabolites play a crucial role in modulating the effects of inflammatory cytokines on cardiovascular health.

Despite the strengths of our study, several limitations should be acknowledged. First, although MR is a powerful tool for inferring causal relationships, it is not immune to biases such as pleiotropy, where genetic variants influence multiple traits through different pathways [29]. While we employed sensitivity analyses, including MR-Egger and weighted median, to account for pleiotropy, residual confounding cannot be entirely ruled out. Second, our study population consisted primarily of individuals of European descent, which may limit the generalizability of our findings to other ethnic groups. Future studies should aim to replicate these results in more diverse populations to ensure the robustness of the associations. Additionally, while we identified several ICs and metabolites that are causally linked to arrhythmia, the exact biological mechanisms through which these factors influence arrhythmogenesis remain unclear. Further experimental studies are needed to validate these findings and elucidate the underlying molecular pathways. Finally, the use of summary-level data in MR analyses precludes the investigation of individual-level variability in genetic and environmental factors that may also influence arrhythmia risk.

In conclusion, our study provides compelling evidence for the complex interplay between inflammatory cytokines, metabolites, and the development of arrhythmias. The identification of OSM, CXCL11, and FGF5 as significant contributors to arrhythmia risk, as well as the protective effects of IL-6 and CD40L, offers novel insights into the inflammatory and metabolic regulation of cardiac electrophysiology. Moreover, the involvement of metabolites such as sphingomyelins and 1-palmitoyl-2-oleoyl-GPE underscores the importance of lipid metabolism in arrhythmogenesis. These findings suggest that targeting specific inflammatory and metabolic pathways could be a viable therapeutic strategy for reducing arrhythmia risk. Future research should focus on exploring the underlying biological mechanisms and expanding these analyses to more diverse populations.

## Conclusions

The findings reveal a complex interplay between ICs, metabolites, and arrhythmia risk. The mediation effect of 1-palmitoyl-2-oleoyl-GPE (16:0/18:1) in the FGF5-arrhythmia causal pathway suggests metabolic regulation’s critical role in the pathogenesis of arrhythmia. Using advanced genetic approaches like Mendelian Randomization, the research provides insights into how these biological processes may contribute to cardiovascular disease. While progress has been made in identifying potential links, further research is needed to explore the underlying molecular mechanisms and confirm causality across diverse populations. Future studies should integrate multi-omics approaches and include different ethnic groups to ensure broader applicability.

## Data Availability

All relevant data are within the manuscript and its Supporting Information files.

## Acknowledgements

We thank the GWAS website and the Finnish database for providing valuable information.

## Author Contributions

Yu-fei Xie and Ling-hui Tang contributed equally to this work. They were responsible for the design, conception of the study and data analysis. Zhi-yu Zeng and Feng Huang are the corresponding authors. They supervised the overall project, provided intellectual input, secured funding, and revised the manuscript critically for important intellectual content.

## Availability of data and materials

The datasets presented in this study can be found in online repositories. The names of the repository/repositories and accession number(s) can be found in the article/Additional Material.

## Ethics approval and consent to participate

This study relied on publicly accessible summary statistics from previously published research and consortia. Each original study involved in this research had received ethical clearance from their respective review boards, and all participants had given informed consent. Notably, this research did not involve the use of individual-level data, thereby eliminating the need for new ethical review board approval.

